# Postoperative Atrial Fibrillation After Coronary Artery Bypass Grafting and Its Association with Length of Stay, Discharge Disposition, and 90-Day Outcomes

**DOI:** 10.64898/2026.06.23.26356270

**Authors:** Leonardo Alexis Almaguer Gongora, Myrna Eliann Reinhardt, Manuel Jimenez Jimenez, Luis Enrique Remedios Carbonell, Priya Mohan, Daniel Padron, Juliette Camejo, Carlos Acosta-Batista, Bernardo Reyes

## Abstract

**Background:** Postoperative atrial fibrillation (POAF) is a frequent complication following coronary artery bypass grafting (CABG) and is associated with increased acute morbidity and resource utilization. However, its independent role in driving post-discharge adverse events in contemporary practice remains debated.

**Objective:** To evaluate the association between POAF and short-term outcomes after CABG, and to utilize empirical Bayesian risk updating to stratify 90-day post-discharge vulnerabilities.

**Methods:** A retrospective cohort analysis of 4,684 adult patients who underwent isolated CABG in Florida between January 1, 2021, and June 30, 2024, was conducted, excluding those with documented preoperative AFib. We employed multivariable negative binomial and logistic regression models to assess length of stay (LOS), discharge disposition, 90-day readmission, and 90-day composite complications. Additionally, a Bayesian Beta-Binomial conjugate model with an objective Jeffreys Prior was utilized to estimate the posterior probabilities of adverse outcomes across key clinical phenotypes.

**Results:** POAF occurred in 355 patients (7.58%). Multivariable analysis demonstrated a 30% relative increase in expected LOS (IRR 1.30, 95% CI [1.23–1.36], P < .001) and 33% higher odds of facility discharge (OR 1.33, 95% CI [1.03–1.72], P = .030) for patients with POAF. However, POAF was not independently associated with 90-day readmission (OR 1.25, P = .063) or composite complications (OR 1.20, P = .118). Chronic heart failure (CHF) emerged as the dominant predictor. Bayesian risk updating revealed that while the baseline posterior probability for a 90-day complication was 27.2%, the synergistic presence of both POAF and CHF radically shifted this posterior risk to 42.6% (Probability of Direction > 0.999 vs. baseline).

**Conclusions:** POAF prolongs hospitalization and drives non-home discharges, but it does not independently dictate 90-day morbidity. Bayesian stratification demonstrates that post-discharge outcomes are predominantly driven by underlying chronic conditions. Effective reduction of readmissions requires robust transition-of-care frameworks, empowering primary care clinicians to aggressively optimize heart failure and metabolic disease rather than focusing solely on the acute surgical arrhythmic event.

## Introduction

Postoperative atrial fibrillation (POAF) is traditionally reported to occur in approximately 20% to 40% of patients following coronary artery bypass grafting (CABG) when capturing all transient, self-limiting arrhythmic episodes via continuous telemetry [1–3]. However, clinically significant POAF—defined as episodes severe, symptomatic, or prolonged enough to warrant formal administrative coding and targeted management—represents a distinct, higher-risk phenotype. While POAF has been broadly linked to longer hospital stays, increased healthcare costs, stroke, and early mortality [4–6], it remains a subject of ongoing debate whether these adverse post-discharge outcomes are driven independently by the arrhythmia itself, or if POAF primarily acts as a hospital-based biomarker reflecting underlying patient vulnerability.

Recent studies suggest that improvements in perioperative rhythm monitoring, beta-blocker use, and anticoagulation strategies may have attenuated the long-term consequences historically attributed to POAF [7–9]. Additionally, many prior studies were conducted in earlier surgical eras or lacked adjustment for contemporary comorbidity burdens and modern discharge pharmacotherapy. Consequently, a major challenge during the critical “transitions of care” period is providing primary care and outpatient clinicians with precise tools to identify which patients require aggressive post-discharge monitoring.

Using a large, modern, multicenter cohort of CABG patients, we sought to evaluate the association between clinically significant POAF and (1) hospital length of stay, (2) discharge disposition, (3) 90-day readmission, and (4) 90-day postoperative complications. Furthermore, to translate population-level models into actionable clinical metrics, we employed an empirical Bayesian risk-updating framework. We hypothesized that while POAF strongly dictates acute in-hospital outcomes and non-home discharge dispositions, adverse 90-day post-discharge events are not independently driven by the arrhythmia, but rather by the synergistic presence of underlying chronic cardiovascular and metabolic disease.

## Methods

### Study Design and Population

This retrospective observational cohort study included adult patients (≥18 years) who underwent CABG between January 1, 2021, and June 30, 2024, across multiple hospitals in Florida. Patients with evidence of atrial fibrillation prior to surgery—including diagnosis codes, nursing documentation, or home antiarrhythmic therapy—were excluded. Additional exclusions included missing key covariates, in-hospital mortality, discharge against medical advice, incarceration, and duplicate encounters. The final analytic cohort consisted of 4,684 patients.

### Exposure and Outcomes

The primary exposure was clinically significant POAF, defined by the presence of new-onset atrial fibrillation diagnosis codes (ICD-10 I48) during the index CABG hospitalization in patients without preoperative atrial fibrillation. This administrative definition specifically captures arrhythmic events severe or prolonged enough to require formal clinical documentation and management.

The primary outcome was hospital length of stay (LOS). Secondary outcomes included discharge disposition (home vs. facility), 90-day readmission, and a composite of 90-day postoperative complications (ischemic stroke, hemorrhagic stroke, infection, gastrointestinal bleeding, or acute coronary syndrome).

### Covariates

Models adjusted for age group, sex, race, body mass index category, diabetes, peripheral arterial disease, chronic obstructive pulmonary disease (COPD), chronic heart failure (CHF), chronic kidney disease, and smoking status. Readmission models were additionally adjusted for discharge medications, including beta-blockers, antiarrhythmics, anticoagulants, antiplatelets, ACE inhibitors, ARBs, and calcium channel blockers.

### Statistical Analysis

Negative binomial regression was used to model LOS due to overdispersion. Binary logistic regression models were applied for all dichotomous outcomes. Model discrimination and calibration were assessed using c-statistics and Hosmer–Lemeshow tests where applicable. All analyses were performed using SAS version 9.4 (SAS Institute Inc).

To translate the adjusted population-level models into actionable clinical metrics, absolute risk and phenotype disparities were modeled through empirical Bayesian inference. We employed a Bayesian Beta-Binomial model for the comparison of proportions to estimate the posterior distributions of 90-day readmissions and complications across key clinical phenotypes (POAF and Congestive Heart Failure [CHF]). We utilized an objective, non-informative Jeffreys Prior (Beta(0.5, 0.5)) for all parameters. This choice ensures that the posterior distributions are driven almost entirely by the empirical data structure while maintaining mathematical invariance to reparameterization. The primary Bayesian metric reported is the Probability of Direction (pd), representing the exact probability that the observed disparity between any two phenotypic groups is strictly greater than zero.

### Ethics Statement

The requirement for formal Institutional Review Board (IRB) approval and informed consent was waived for this project. This study involved a retrospective analysis of pre-existing, fully de-identified administrative clinical data. Because the research relied exclusively on anonymized records and did not involve interaction with human subjects or the use of identifiable protected health information (PHI), it was deemed exempt from ethical review in accordance with the regulatory criteria of the U.S. Department of Health and Human Services (45 CFR 46.104). All data extraction, handling, and analysis were conducted in strict compliance with the Health Insurance Portability and Accountability Act (HIPAA) Privacy Rule.

## Results

### Study Population

Between January 2021 and June 2024, a total cohort of patients undergoing coronary artery bypass grafting was identified. After exclusion of patients with pre-existing atrial fibrillation, missing key covariates, non-routine discharges, or in-hospital mortality, the final analytic cohort consisted of 4,684 patients (Figure 1). Of these, 355 patients (7.6%) developed postoperative atrial fibrillation (POAF) during the index hospitalization.

**Figure 1.**
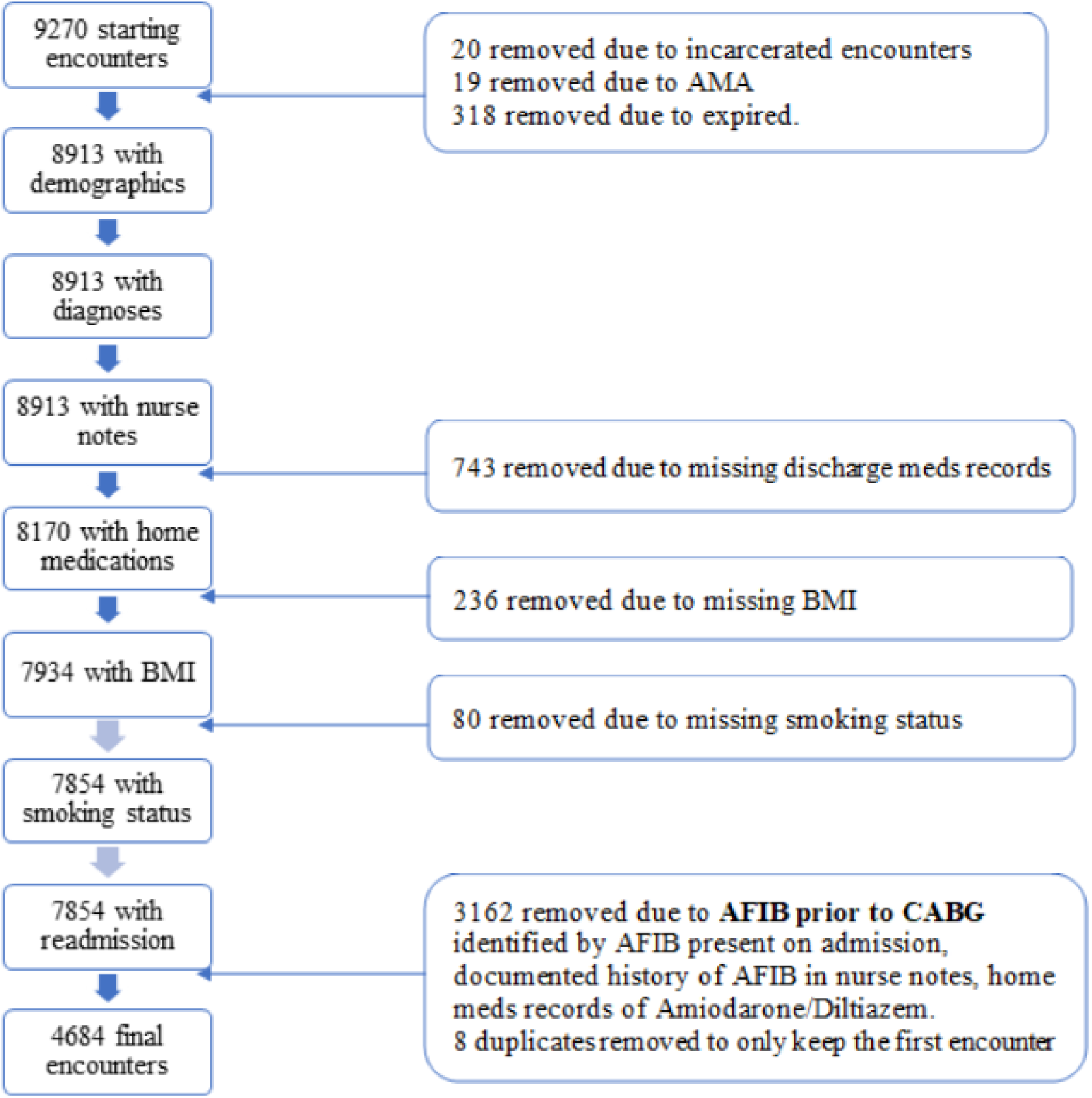
Study Cohort Selection Flow Diagram. Flow diagram illustrating inclusion and exclusion criteria for the study cohort. Adult patients undergoing coronary artery bypass grafting between January 2021 and June 2024 were included. Patients with pre-existing atrial fibrillation, missing key covariates, in-hospital mortality, or non-routine discharges were excluded, yielding a final analytic cohort of 4,684 patients.

Baseline demographic and clinical characteristics stratified by POAF status are shown in Table 1. Patients who developed POAF were older, with a higher proportion aged ≥80 years, and had a greater prevalence of chronic heart failure and chronic kidney disease. Sex distribution and body mass index categories were similar between groups.

**Table 1.**
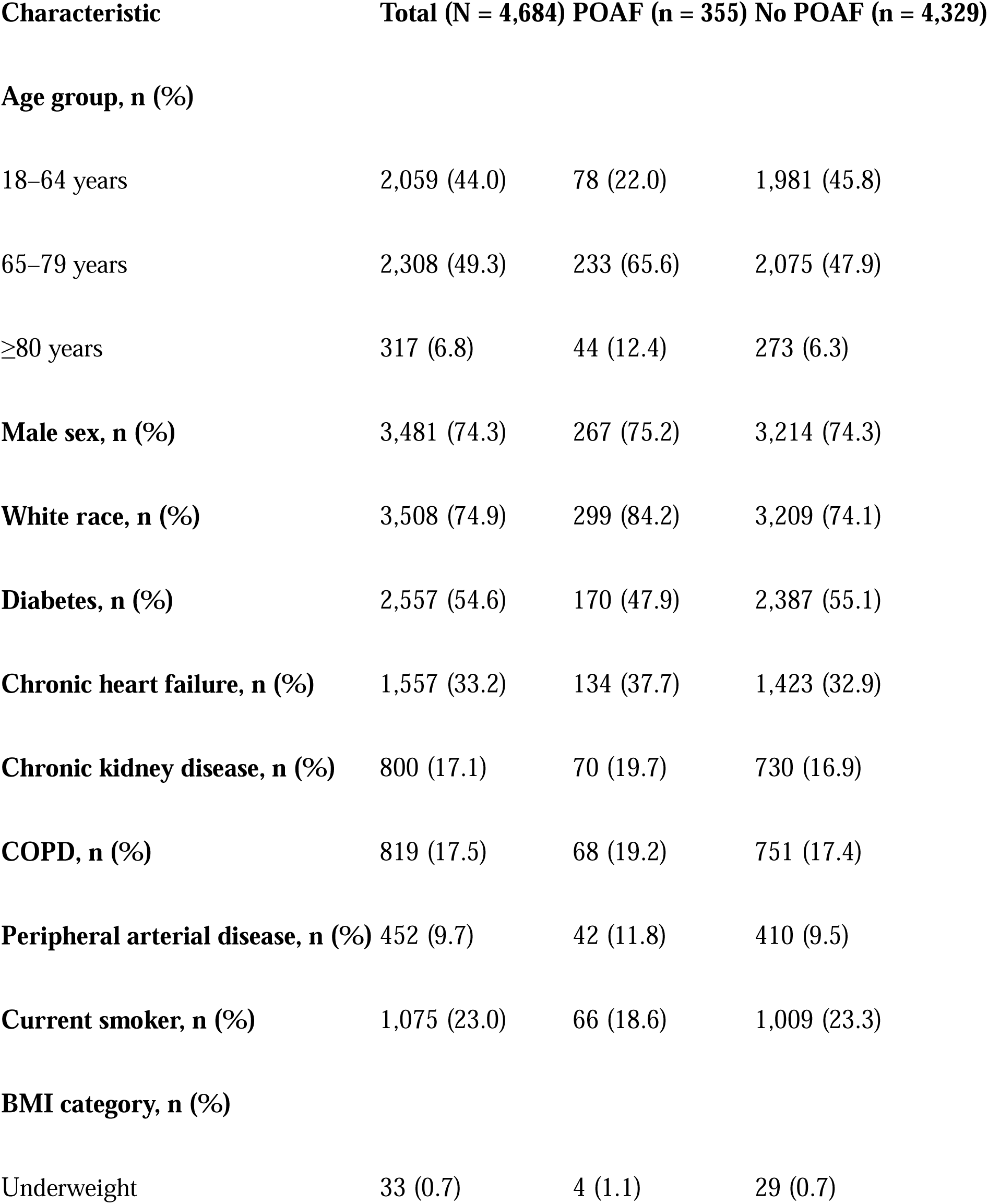
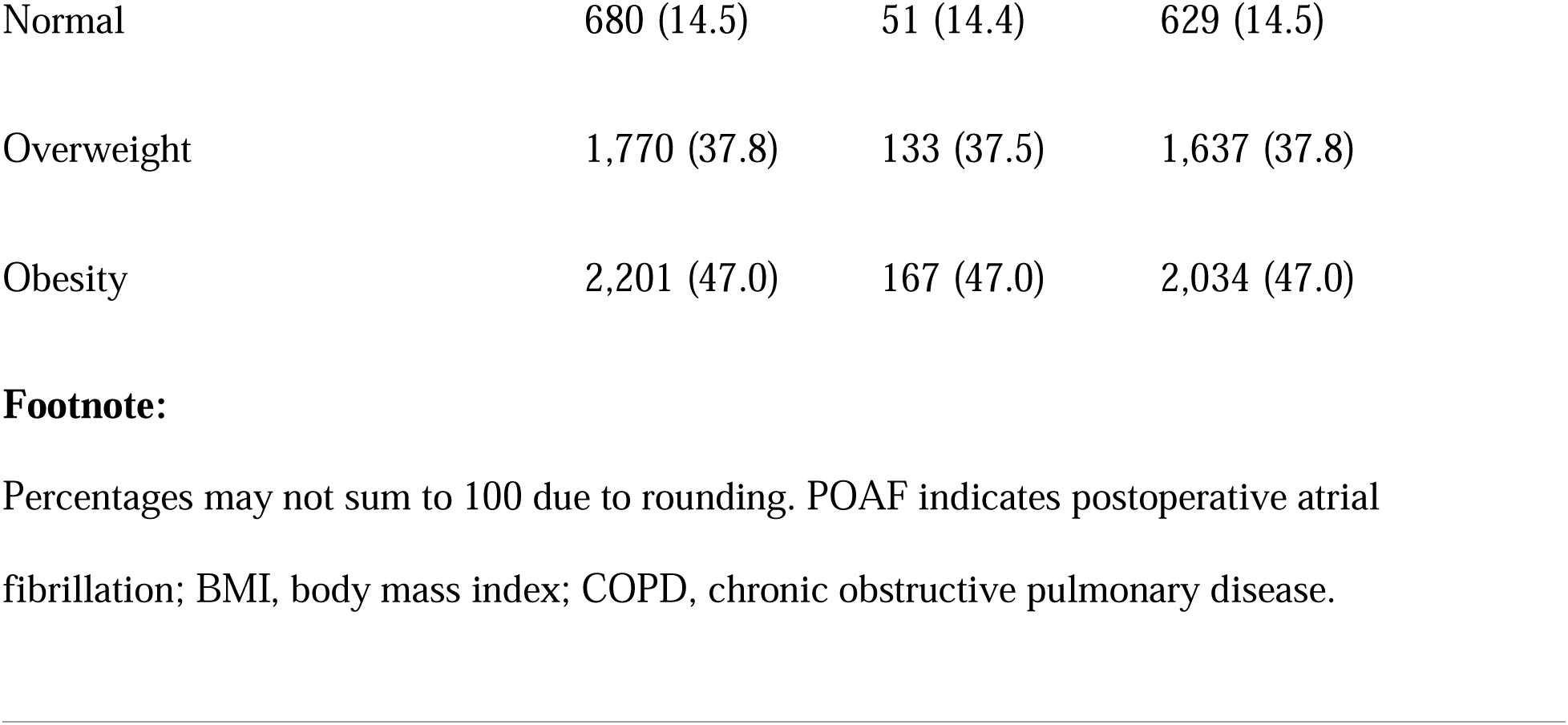
Baseline Characteristics of Patients Undergoing CABG, Stratified by Postoperative Atrial Fibrillation.

### Hospital Length of Stay

Median hospital length of stay was longer among patients with POAF compared with those without POAF (10 [IQR 7–14] vs 8 [IQR 6–12] days; Table 2).

**Table 2.** Unadjusted Clinical Outcomes Stratified by Postoperative Atrial Fibrillation.

| **Outcome** | **Total (N =**<br>**4,684)** | **POAF (n =**<br>**355)** | **No POAF (n =**<br>**4,329)** |
| --- | --- | --- | --- |
| Length of stay, days, median (IQR) | 9 (6–12) | 10 (7–14) | 8 (6–12) |
| Discharge to facility, n (%) | 1,002 (21.4) | 109 (30.7) | 893 (20.6) |
| 90-day readmission, n (%) | 2,195 (46.9) | 188 (53.0) | 2,007 (46.4) |
| 90-day postoperative complications*, n (%) | 1,441 (30.8) | 125 (35.2) | 1,316 (30.4) |

\*Composite of ischemic stroke, hemorrhagic stroke, infection, gastrointestinal bleeding, or acute coronary syndrome.

After multivariable adjustment using negative binomial regression, POAF was independently associated with increased hospital length of stay (IRR 1.30, 95% CI 1.23–1.36; P<0.001). This corresponded to a 30% relative increase in expected length of stay compared with patients without POAF (Table 3).

**Table 3.**
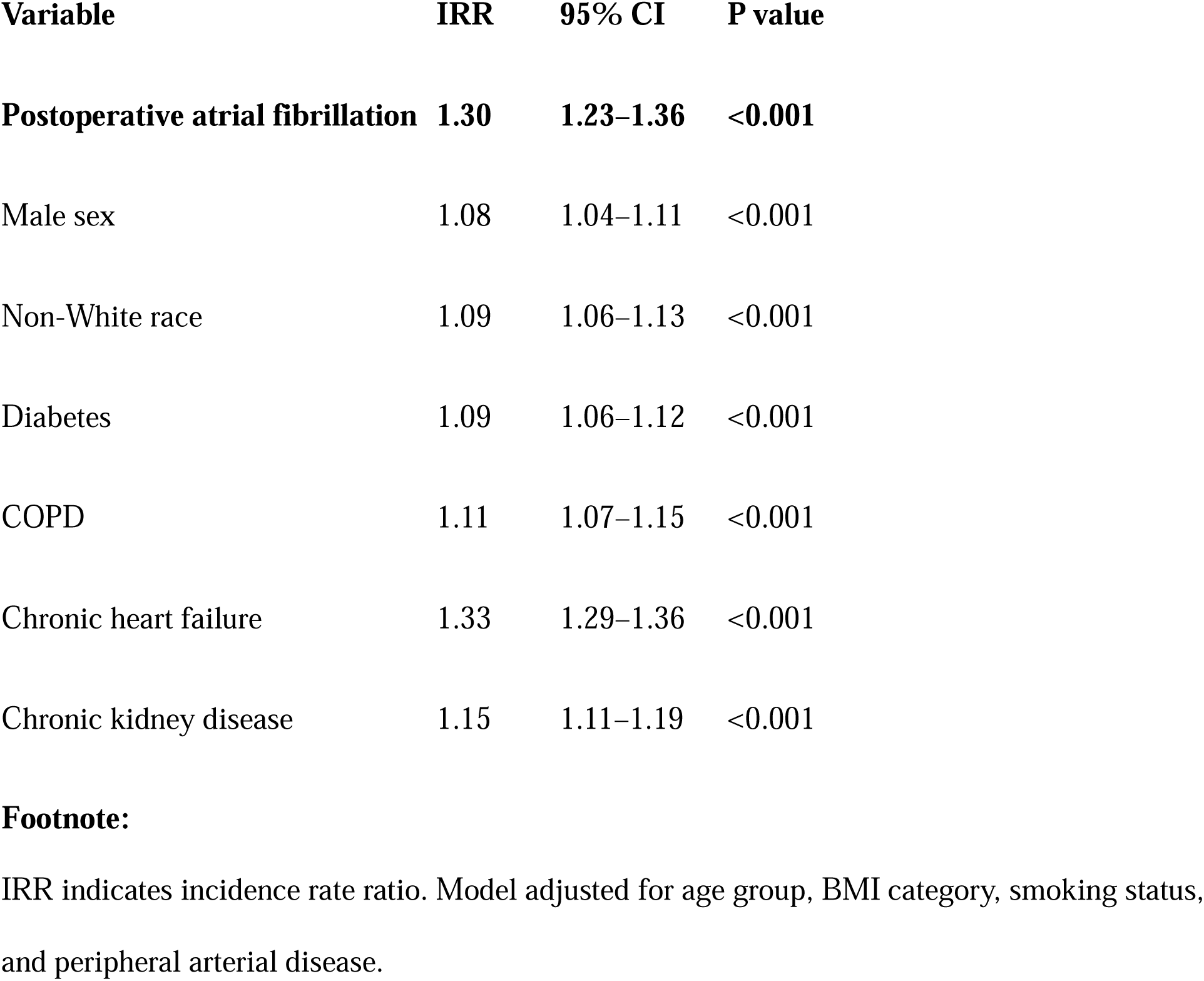
Multivariable Negative Binomial Regression for Length of Stay.

| **Variable** | **IRR** | **95% CI** | **P value** |
| --- | --- | --- | --- |
| **Postoperative atrial fibrillation** | **1.30** | **1.23–1.36** | **<0.001** |
| Male sex | 1.08 | 1.04–1.11 | <0.001 |
| Non-White race | 1.09 | 1.06–1.13 | <0.001 |
| Diabetes | 1.09 | 1.06–1.12 | <0.001 |
| COPD | 1.11 | 1.07–1.15 | <0.001 |
| Chronic heart failure | 1.33 | 1.29–1.36 | <0.001 |
| Chronic kidney disease | 1.15 | 1.11–1.19 | <0.001 |

IRR indicates incidence rate ratio. Model adjusted for age group, BMI category, smoking status, and peripheral arterial disease.

Among covariates, chronic heart failure demonstrated the strongest association with prolonged hospitalization (IRR 1.33, 95% CI 1.29–1.36), followed by chronic kidney disease and chronic obstructive pulmonary disease. Current smoking status was not independently associated with length of stay.

These adjusted associations are visually summarized in Figure 2, which demonstrates the relative contribution of POAF and key comorbidities to prolonged hospitalization.

**Figure 2.**
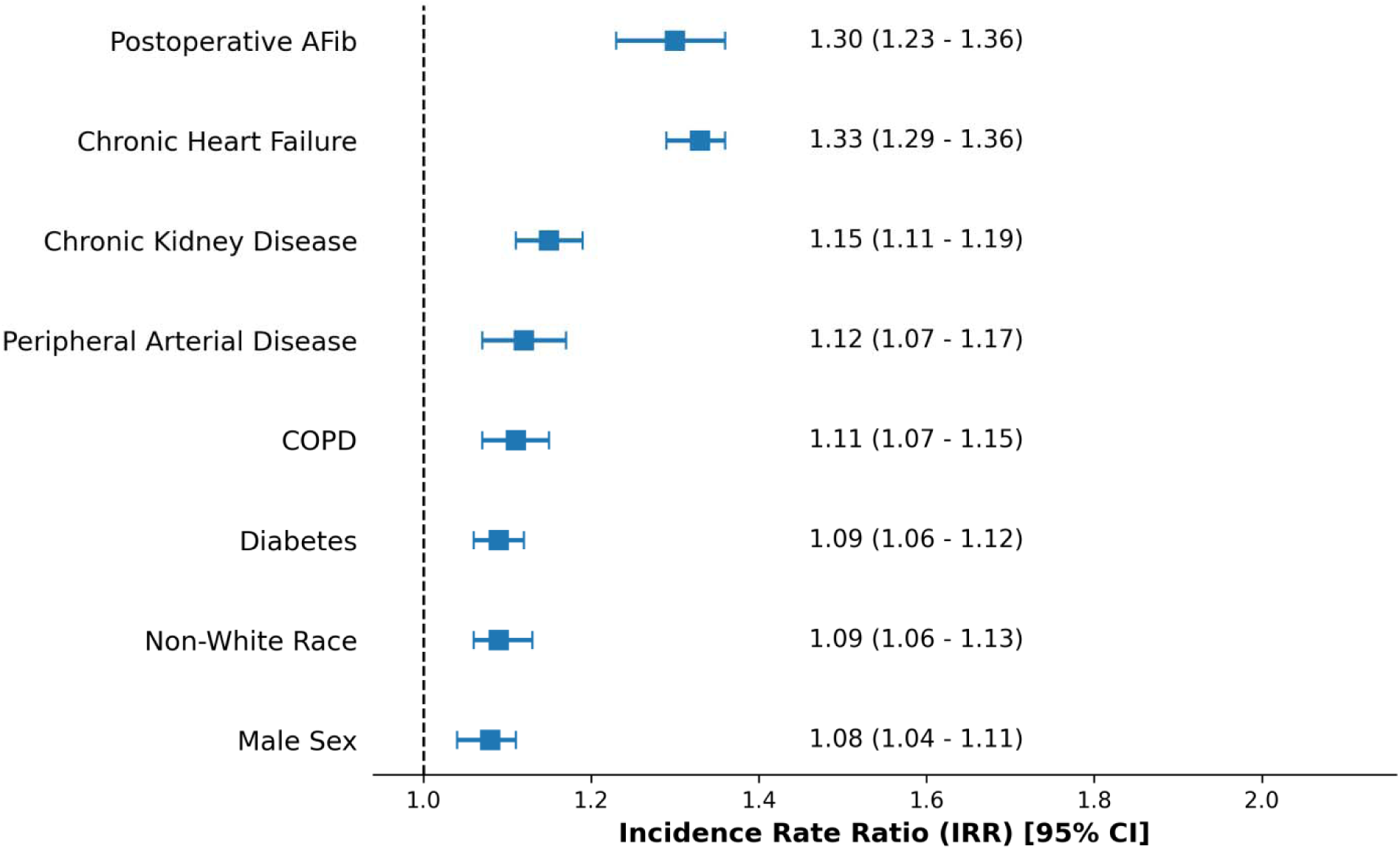
Adjusted Association with Hospital Length of Stay. Forest plot demonstrating adjusted incidence rate ratios for hospital length of stay derived from multivariable negative binomial regression. Postoperative atrial fibrillation was independently associated with a 30% increase in hospital length of stay after adjusting for demographics and comorbidities.

### Discharge Disposition

Overall, 21.4% of patients were discharged to a facility rather than home. Discharge to a facility occurred more frequently among patients with POAF than among those without POAF (30.7% vs 20.6%; Table 2).

In multivariable logistic regression, POAF was independently associated with increased odds of discharge to a facility (OR 1.33, 95% CI 1.03–1.72; P=0.030) after adjustment for demographics and comorbidities (Table 4). Advanced age and chronic heart failure were the strongest predictors of non-home discharge, while male sex was associated with lower odds of facility discharge. The adjusted odds ratios for discharge disposition are illustrated in Figure 3.

**Figure 3.**
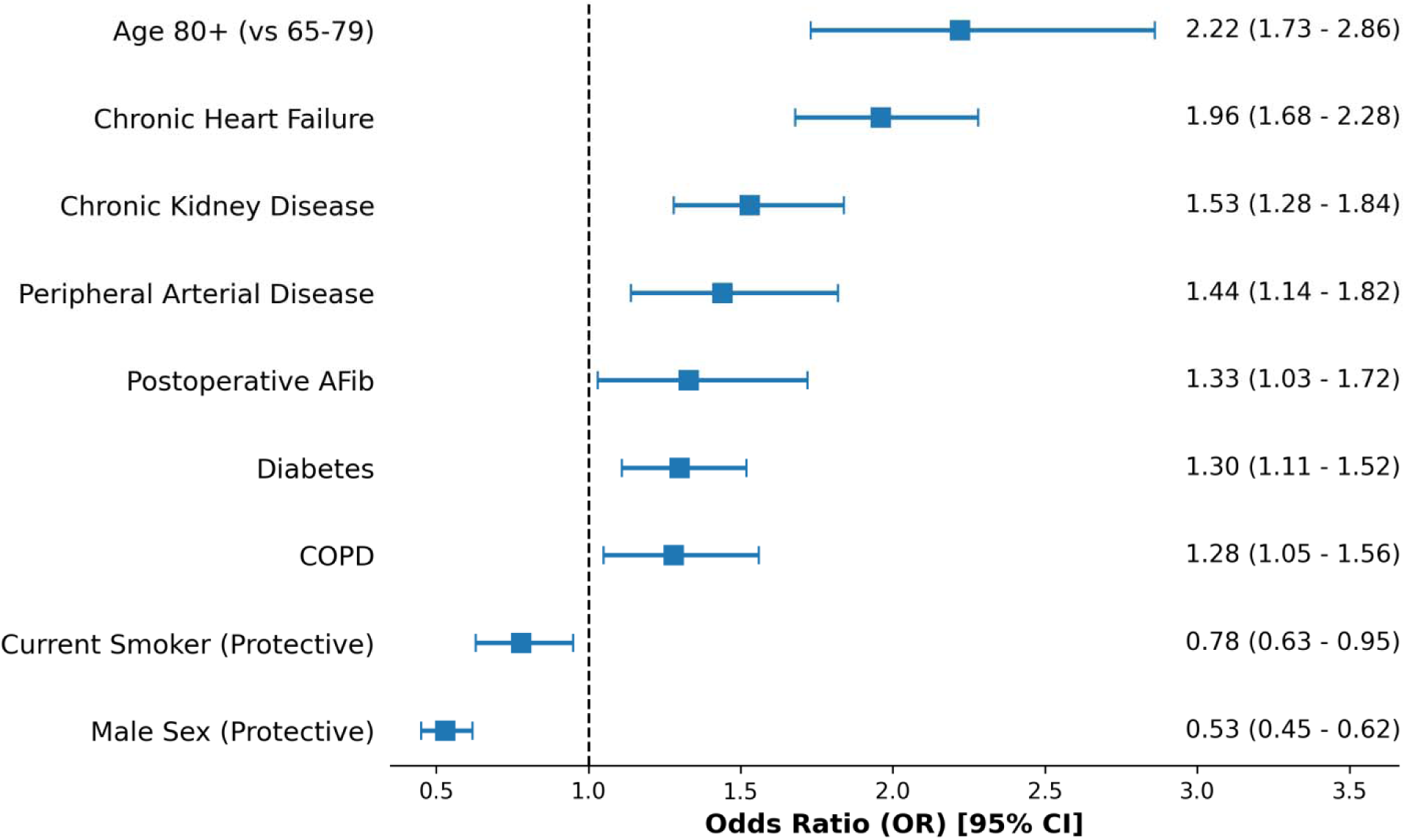
Adjusted Odds of Discharge to Facility. Forest plot of adjusted odds ratios for discharge to a facility rather than home. Postoperative atrial fibrillation, advanced age, and chronic heart failure were associated with increased likelihood of non-home discharge.

**Table 4.** Multivariable Logistic Regression for Discharge to Facility.

| **Variable** | **OR** | **95% CI** | **P value** |
| --- | --- | --- | --- |
| **Postoperative atrial fibrillation** | **1.33** | **1.03–1.72** | **0.030** |
| Age $\geq 80$ vs 65–79 years | 2.22 | 1.73–2.86 | <0.001 |
| Male sex | 0.53 | 0.45–0.62 | <0.001 |
| Diabetes | 1.30 | 1.11–1.52 | 0.001 |
| Chronic heart failure | 1.96 | 1.68–2.28 | <0.001 |
| Chronic kidney disease | 1.53 | 1.28–1.84 | <0.001 |

### Ninety-Day Readmission

Within 90 days of discharge, 46.9% of patients experienced at least one readmission. Readmission occurred in 53.0% of patients with POAF and 46.4% of patients without POAF (Table 2).

After adjustment for demographics, comorbidities, and discharge medications, POAF was not independently associated with 90-day readmission (OR 1.25, 95% CI 0.99–1.59; P=0.063; Table 5). Chronic heart failure was the only comorbidity independently associated with readmission risk (OR 1.28, 95% CI 1.13–1.46), while male sex was associated with lower odds of readmission.

**Table 5.** Multivariable Logistic Regression for 90-Day Readmission.

| **Variable** | **OR** | **95% CI** | **P value** |
| --- | --- | --- | --- |
| Postoperative atrial fibrillation | 1.25 | 0.99–1.59 | 0.063 |
| Male sex | 0.84 | 0.74–0.96 | 0.011 |
| Chronic heart failure | 1.28 | 1.13–1.46 | <0.001 |

### Ninety-Day Postoperative Complications

The composite endpoint of postoperative complications within 90 days occurred in 30.8% of patients overall (Table 2). Although complication rates were numerically higher among patients with POAF, POAF was not independently associated with postoperative complications after multivariable adjustment (OR 1.20, 95% CI 0.95–1.52; P=0.118; Table 6), however, diabetes, chronic obstructive pulmonary disease, and chronic heart failure were independently associated with increased odds of postoperative complications, whereas male sex was associated with lower risk.

**Table 6.**
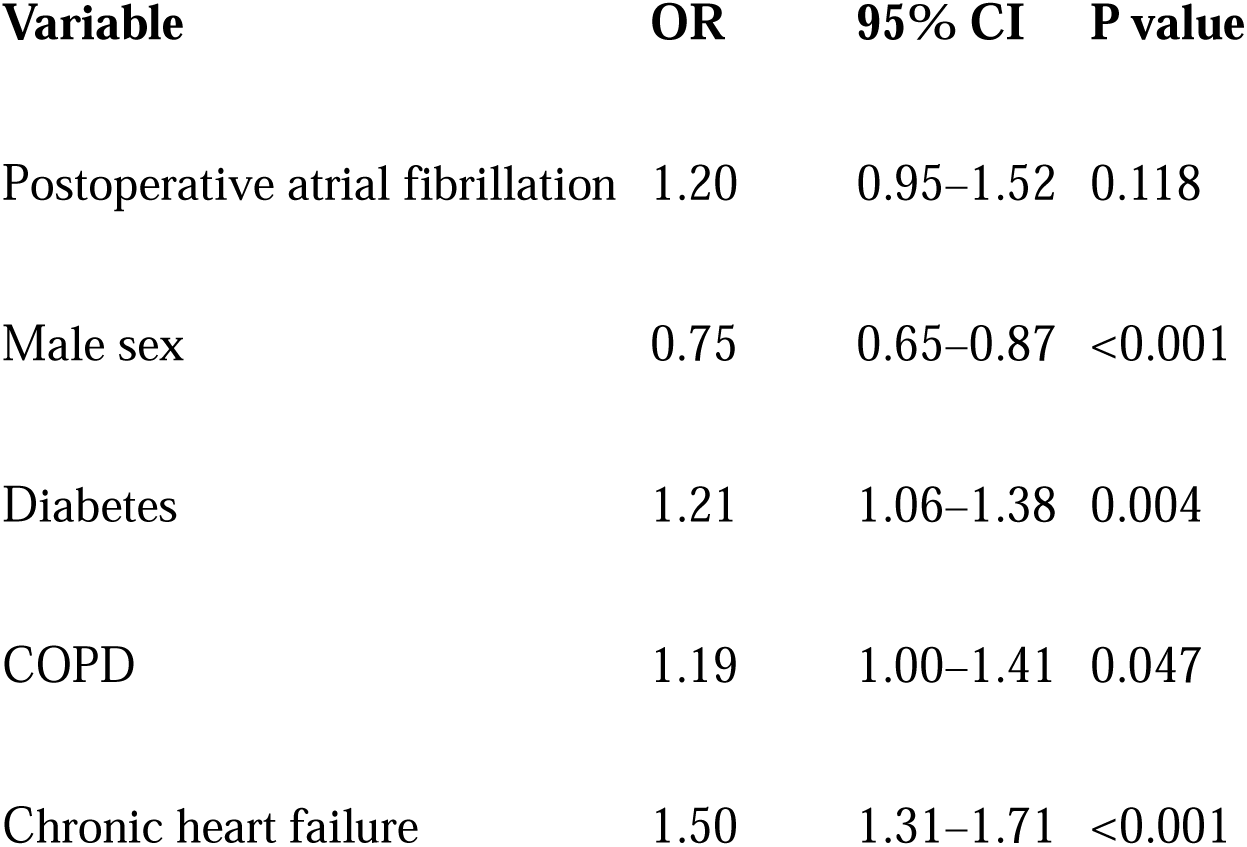
Multivariable Logistic Regression for 90-Day Postoperative Complications.

| **Variable** | **OR** | **95% CI** | **P value** |
| --- | --- | --- | --- |
| Postoperative atrial fibrillation | 1.20 | 0.95–1.52 | 0.118 |
| Male sex | 0.75 | 0.65–0.87 | <0.001 |
| Diabetes | 1.21 | 1.06–1.38 | 0.004 |
| COPD | 1.19 | 1.00–1.41 | 0.047 |
| Chronic heart failure | 1.50 | 1.31–1.71 | <0.001 |

### Bayesian Posterior Distributions and Probability of Direction

To further stratify clinical risk based on the dominant predictor (CHF) identified in our multivariable models, posterior distributions were estimated for the prevalence of 90-day outcomes across four clinical phenotypes. For 90-day readmissions, the baseline phenotype (No POAF, No CHF) demonstrated a posterior mean of 44.29% (95% Credible Interval [CrI]: 42.49% - 46.10%). The presence of an isolated POAF episode increased the posterior mean to 48.87%, yielding a Probability of Direction (pd) of 0.906 against the baseline. However, underlying CHF alone drove the posterior readmission probability to 50.60% (pd > 0.999 vs. baseline). The synergistic combination of POAF and CHF resulted in the highest posterior readmission risk (59.63%), demonstrating a 97.8% exact probability (pd = 0.978) of being strictly greater than the risk of CHF alone.

Similar Bayesian updates were observed for 90-day severe complications. The baseline posterior mean was 27.19% (95% CrI: 25.59% - 28.82%). Isolated POAF marginally updated this to 30.86% (pd = 0.874 vs. baseline). In contrast, the combination of POAF and CHF radically shifted the posterior distribution to 42.59% (95% CrI: 34.40% - 50.99%), representing a near-certain disparity (pd > 0.999) compared to the baseline phenotype (Table 7).

**Table 7.** Bayesian Posterior Distributions for 90-Day Outcomes Stratified by POAF and Congestive Heart Failure.

| **Clinical Phenotype** | **Readmission Posterior Mean**<br>**(95% CrI)** | **Complication Posterior Mean**<br>**(95% CrI)** |
| --- | --- | --- |
| **No POAF / No CHF** | 44.29% (42.49% – 46.10%) | 27.19% (25.59% – 28.82%) |
| **POAF / No CHF** | 48.87% (42.33% – 55.44%) | 30.86% (24.96% – 37.07%) |
| **No POAF / CHF** | 50.60% (48.00% – 53.19%) | 36.97% (34.48% – 39.50%) |
| **POAF / CHF** | 59.63% (51.26% – 67.73%) | 42.59% (34.40% – 50.99%) |

Note: Posterior means and 95% Credible Intervals (CrI) were estimated using a Beta-Binomial conjugate model with an objective Jeffreys Prior (Beta(0.5, 0.5)).

## Discussion

In this contemporary, multicenter CABG cohort, POAF was strongly associated with increased hospital LOS and greater need for post-acute care but was not independently associated with 90-day readmission or postoperative complications. These findings suggest that POAF primarily reflects acute postoperative complexity rather than driving downstream adverse events.

Our results align with recent studies demonstrating that while POAF increases short-term resource utilization, its impact on longer-term outcomes may be attenuated by modern perioperative management [10–12]. The lack of association between POAF and readmission contrasts with earlier studies conducted before the widespread adoption of beta-blockers and structured discharge planning [13–15]. This shift highlights the evolving clinical significance of POAF in the contemporary era.

Crucially, congestive heart failure (CHF) consistently emerged as the dominant predictor across all post-discharge outcomes, underscoring the central role of underlying cardiac dysfunction.

From a clinical decision-making standpoint, these findings highlight the utility of a Bayesian Beta-Binomial framework in transitions of care. The data demonstrate that while POAF is a primary driver of non-home discharges (facility rehabilitation)—exposing patients to early risks of deconditioning—our posterior probability estimates reveal that the true drivers of 90-day readmissions and severe postoperative complications are underlying chronic conditions.

For primary care physicians managing these patients post-discharge, the occurrence of POAF should serve as an early, hospital-based biomarker for systemic vulnerability. While an isolated POAF episode updates the 90-day complication risk marginally from the baseline (pd = 0.874), its presence alongside CHF synergistically shifts the posterior probability of a severe complication to 42.59%. This signals the absolute necessity for an aggressive, multidisciplinary care coordination model upon discharge, prioritizing the optimization of volume status and metabolic control over isolated rhythm monitoring.

### Limitations

This study is observational and cannot establish causality. POAF was identified using administrative coding (ICD-10) and may be subject to misclassification, potentially capturing only the most clinically significant events. Residual confounding remains possible despite extensive multivariable adjustment. Furthermore, readmissions outside the participating hospital system may not have been captured. Finally, while our empirical Bayesian framework utilized an objective Jeffreys Prior to minimize bias, the posterior probabilities are inherently dependent on the specific phenotypic definitions and cohort prevalence observed within this regional health system.

### Conclusions

Among patients undergoing CABG in a modern healthcare system, POAF is associated with longer hospitalization and an increased likelihood of discharge to a rehabilitation facility, but it does not independently drive 90-day readmission or systemic postoperative complications. Our Bayesian risk stratification highlights that post-discharge preventive strategies must evolve. True reduction in 90-day morbidity requires a robust transition-of-care framework that empowers outpatient clinicians to aggressively optimize underlying heart failure and metabolic disease in these highly vulnerable high-risk phenotypes, rather than focusing solely on the acute surgical arrhythmic event.

## Data Availability

All data produced in the present study are available upon reasonable request to the authors

## Glossary of Abbreviations

AF: Atrial fibrillation
POAF: Postoperative atrial fibrillation
CABG: Coronary artery bypass grafting
LOS: Length of stay
OR: Odds ratio
IRR: Incidence rate ratio
CI: Confidence interval
CHF: Congestive heart failure
COPD: Chronic obstructive pulmonary disease
CKD: Chronic kidney disease
PAD: Peripheral artery disease
BMI: Body mass index

## References

[1] Herrmann FEM, Jeppsson A, Kirov H, et al. Long-Term Continuous Monitoring of New-Onset Atrial Fibrillation After Coronary Artery Bypass Grafting. JAMA. 2025;334(20):1827–1835. doi:10.1001/jama.2025.14891

[2] Gaudino M, Di Franco A, Rong LQ, Piccini J, Mack M. Postoperative atrial fibrillation: from mechanisms to treatment. Eur Heart J. 2023;44(12):1020–1039. doi:10.1093/eurheartj/ehad019

[3] Zhou JY, Zhang JL, Xi L, et al. Risk Factors of Postoperative Atrial Fibrillation After Isolated Coronary Artery Bypass Grafting Surgery in the Recent 10 Years: Clinical Analysis of 6229 Patients. Clin Cardiol. 2024;47(10):e24335. doi:10.1002/clc.24335

[4] van de Kar MRD, van Brakel TJ, Van’t Veer M, et al. Anticoagulation for post-operative atrial fibrillation after isolated coronary artery bypass grafting: a meta-analysis. Eur Heart J. 2024;45(29):2620–2630. doi:10.1093/eurheartj/ehae267.

[5] Taha A, Nielsen SJ, Bergfeldt L, et al. New-Onset Atrial Fibrillation After Coronary Artery Bypass Grafting and Long-Term Outcome: A Population-Based Nationwide Study From the SWEDEHEART Registry. J Am Heart Assoc. 2021;10(1):e017966. doi:10.1161/JAHA.120.017966.

[6] Conen D, Wang MK, Devereaux PJ, et al. New-Onset Perioperative Atrial Fibrillation After Coronary Artery Bypass Grafting and Long-Term Risk of Adverse Events: An Analysis From the CORONARY Trial. J Am Heart Assoc. 2021;10(12):e020426. doi:10.1161/JAHA.120.020426

[7] Makarem A, Paneitz D, Winship T, et al. Pericardiotomy and Amiodarone for Prophylaxis Against Postoperative Atrial Fibrillation in Cardiac Surgery (PAPPA). Ann Thorac Surg. 2025;120(3):461–468. doi:10.1016/j.athoracsur.2025.05.045.

[8] Writing Committee Members, Joglar JA, Chung MK, et al. 2023 ACC/AHA/ACCP/HRS Guideline for the Diagnosis and Management of Atrial Fibrillation: A Report of the American College of Cardiology/American Heart Association Joint Committee on Clinical Practice Guidelines. J Am Coll Cardiol. 2024;83(1):109-279. doi:10.1016/j.jacc.2023.08.017.

[9] Suero OR, Ali AK, Barron LR, Segar MW, Moon MR, Chatterjee S. Postoperative atrial fibrillation (POAF) after cardiac surgery: clinical practice review. J Thorac Dis. 2024;16(2):1503–1520. doi:10.21037/jtd-23-1626.

[10] Cheah DS, Tsai K, Kurpad KP, Khalid Y, Mehta SS. Trends and Outcomes of Anticoagulation for Post-Operative Atrial Fibrillation After Coronary Artery Bypass Graft. Pacing Clin Electrophysiol. 2025;48(12):1444–1450. doi:10.1111/pace.70069.

[11] Rezk M, Taha A, Nielsen SJ, et al. Clinical Course of Postoperative Atrial Fibrillation After Cardiac Surgery and Long-term Outcome. Ann Thorac Surg. 2022;114(6):2209–2215. doi:10.1016/j.athoracsur.2022.03.062

[12] Grant MC, Crisafi C, Alvarez A, et al. Perioperative Care in Cardiac Surgery: A Joint Consensus Statement by the Enhanced Recovery After Surgery (ERAS) Cardiac Society, ERAS International Society, and The Society of Thoracic Surgeons (STS). Ann Thorac Surg. 2024;117(4):669–689. doi:10.1016/j.athoracsur.2023.12.006.

[13] Bianco V, Kilic A, Yousef S, et al. The long-term impact of postoperative atrial fibrillation after cardiac surgery. J Thorac Cardiovasc Surg. 2023;166(4):1073–1083.e10. doi:10.1016/j.jtcvs.2021.10.072.

[14] Shah RM, Zhang Q, Chatterjee S, et al. Incidence, Cost, and Risk Factors for Readmission After Coronary Artery Bypass Grafting. Ann Thorac Surg. 2019;107(6):1782–1789. doi:10.1016/j.athoracsur.2018.10.077.

[15] Khoury H, Sanaiha Y, Rudasill SE, Mardock AL, Sareh S, Benharash P. Readmissions Following Isolated Coronary Artery Bypass Graft Surgery in the United States (from the Nationwide Readmissions Database 2010 to 2014). Am J Cardiol. 2019;124(2):205–210. doi:10.1016/j.amjcard.2019.04.018

